# Factors that influence the implementation of digital health in rural, regional, and remote Australia: Protocol for an overview of reviews

**DOI:** 10.1101/2024.11.13.24317285

**Authors:** Michelle A. Krahe, Stephanie Baker, Leanna Woods, Sarah L. Larkins

## Abstract

**Background:** Although digital transformation holds significant importance for the delivery of health care services, its application in rural, regional, and remote (RRR) settings presents unique challenges which are less comprehensively understood. This review aims to consolidate the available evidence from existing systematic and scoping reviews related to the implementation of digital health in RRR Australia, to examine the health, innovation and implementation outcomes, factors that influence implementation, and the design constructs specific to these settings.

**Methods:** This overview of systematic and scoping reviews will follow the guidelines of the Cochrane Overview of Reviews. Qualitative, quantitative, and mixed-methods reviews of studies focusing on the implementation of digital health in RRR Australia will be in included. We will search the following databases: Cochrane Library, PubMed, SCOPUS, CINAHL, and Web of Science. We will include studies published in English from the inception of the database until May 01, 2024. Two reviewers will conduct all steps independently and a third reviewer will determine and resolve discrepancies. Included reviews will be appraised using the AMSTAR-2 checklist. A theory-based coding of barriers and enablers using the CFIR 2.0 will be applied and a narrative summary of the findings, along with tables and diagrams aligned with the objective and scope of the review will be presented.

**Discussion:** We expect this overview of reviews to provide current evidence and key insights about the factors that inhibit or promote the implementation of digital health in RRR Australia. Findings will orient the selection and adaptation of implementation process strategies tailored to digital health in RRR Australia. We will publish our results in a peer-reviewed journal.

**Systematic review registration:** PROSPERO CRD42024512742

## Introduction

In Australia, seven million people (∼29% of the population) live in rural, regional and remote (RRR) areas, accounting for almost 95% of Australia’s total land mass (1). This means that people are highly distributed over a large area and live in isolation from essential health services. In addition, the time and cost associated with transportation to health care or major medical centres, means that people living in RRR Australia experience delays in preventative and primary health care (2–4). As a result, they experience poorer health outcomes which include higher rates of arthritis, mental and behavioural conditions, chronic obstructive pulmonary disease, diabetes, and lung cancer, and have a mortality rate of 1.3 times higher, compared with all of Australia (5). The delivery of health care in RRR locations is further challenged by the long distances between population centres, persistent health workforce shortages and high turnover rates (6, 7). For example, in 2023, only 1.2% of all registered and employed clinicians were located in remote locations (8), and the higher burden of disease in rural Australia represents $27.5b loss in economic contributions (9). The challenge is providing health care and services in RRR Australia is acute.

One solution that offers real opportunity for people living in RRR Australia is the adoption of digital health innovations (DHIs) (10, 11). These innovations, such as telehealth, mobile health or patient portals leverage the use of digital and data to improve how health services are accessed, used, and delivered to help people live healthier lives (12). Despite the rapid progress in digital health, much of its development remains concentrated in major urban centres, with limited attention given to its application in RRR Australia (4). Given the existing digital divide between metropolitan and RRR areas, there is an urgent need for evidence on optimal strategies for implementing DHI in these areas (12, 13).

Methodologies derived from implementation science can offer valuable insights about the factors influencing the implementation of digital health in RRR settings. The Consolidated Framework for Implementation Research (CFIR), for instance, is a meta-theoretical framework providing a repository of standardised implementation-related constructs at the individual, organisational, and external levels that can be applied across the spectrum of implementation research (14). It can serve as a roadmap for exploring the determinants that shape the implementation of digital health. The CFIR presents a comprehensive framework encompassing five domains pertaining to the characteristics of the innovation being implemented, the implementation process itself, the individuals who are involved, and both the inner and outer settings. Using a framework to identify the factors influencing the implementation of digital health within the context of RRR Australia is critical to tailoring effective implementation process strategies that address key barriers and enhance the delivery of DHIs. This approach ensures that implementation strategies are better aligned to service delivery needs, over research and evaluation, which is often considered as an afterthought. However, selecting or adapting the most suitable implementation strategies can be a challenging task given the extensive options available (15). Thus, a review of the current evidence will guide future research and implementation of digital health in RRR settings. An overview of systematic and scoping reviews was chosen to navigate the challenge of collating, assessing, and synthesising evidence from multiple reviews on a specific topic (16, 17).

### Review questions

In this overview of reviews, we aim to consolidate the available evidence from existing quantitative, qualitative, and mixed-methods systematic and scoping reviews related to factors that influence the implementation of DHIs in RRR Australia. To fulfill this objective, the research questions were formulated through collaborative discussion with the research team. The primary question that serves as a guide for the review is:

1. What are the characteristics of digital health innovations implemented in RRR Australia? Additionally, the review will also focus on the following secondary questions:
2. What are the health, innovation and implementation outcomes reported?
3. What are the barriers and enablers to successful implementation of digital health in rural and remote Australia?
4. What are the design considerations specific to the implementation of digital health in RRR Australia?

## Methods

This proposed overview of reviews protocol follows the Cochrane Overviews of Reviews guideline (18) and is reported in accordance with the Preferred Reporting Items for Systematic review and Meta-Analysis Protocols (PRISMA-P) (19). A completed PRISMA-P checklist can be found in Additional file 1. Findings will be reported in accordance with the Preferred Reporting Items for Overviews of Reviews (PRIOR) statement (20). This protocol has been registered with International Prospective Register of Systematic Reviews (PROSPERO CRD42024512742).

### Search strategy

This search strategy combines search terms that cover four concepts: (1) digital health, (2) RRR, (3) implementation, and (4) barriers and enablers (Additional file 2). The initial search was conducted on 01 May 2024. All identified reviews were imported into a reference management system (Endnote V.20) for removal of duplicates (automatic and manual). After excluding duplicates, 513 references are included for screening.

### Key terms and their definitions

#### Patient, consumer, people

In this review we use the term ‘patient’ to refer to a person receiving treatment for a disease or injury and when referring to members in the community involved in services related to health and wellbeing, we use the term ‘consumers’ or ‘people’ as appropriate. The term ‘healthcare provider’ includes individual health professionals and the organisations for whom they work.

#### Digital health

Recognising the difficulty in one all-encompassing definition for digital health, the broad definition as defined by the Australian Institute of Health and Welfare, best represents the field *“… an umbrella term referring to a range of technologies that can be used to treat patients and collect and share a person’s health information. In Australia, digital health has a broad scope, and includes mobile health and applications (such as SMS reminders via mobile messaging, wellness apps, Medicare Online and COVID check-in apps); electronic prescribing; electronic health records (including My Health Record); telehealth and telemedicine; wearable devices (such as fitness trackers and monitors); robotics and artificial intelligence*” (21).

#### Digital health innovation (DHI)

We adopt the Silberman et.al (22) definition of DHI that meet three criteria (1) the DHI falls within one of three digital health categories (digital health, digital medicine, digital therapeutics), (2) the product is designed to change one or more health behaviours, (3) the value of the product is contingent on the degree to which it improves one or more health outcomes (i.e., clinical, quality of care, healthcare services).

#### Implementation process strategies

Strategies for the implementation process aim at the “process” level of the CFIR, concentrating on the effectiveness of teams or individuals responsible for carrying out the activities necessary to select, modify, and incorporate an intervention (23–25). Implementation process strategies are the approaches or methodologies employed to enhance the adoption, implementation, and sustainability of evidence-based interventions (15).

#### Rural, regional, and remote (RRR)

This report will use the term RRR to describe any area outside of Australia’s major cities as according to the Australian Statistical Geography Standard (ASGS) Edition 3 remoteness structure (26). The ASGS divides Australia into five classes of remoteness (major cities, inner regional, outer regional, remote, very remote) which are characterised by relative geographic access to services. We will apply the RRR classification outlined in the paper unless specific locations are provided, in which case we will consult the ASGS.

### Information sources

We will search for reviews published in the following databases: PubMed via MEDLINE, SCOPUS via Elsevier, Web of Science via Clarivate Analytics (1900 to present), CINAHL Complete via EBSCO Host, and the Cochrane Library. Additional strategies to complement our exploration include examining references cited in eligible reviews, searching for authors who have published extensively in the field, and conducting backward/forward citation searches of related systematic reviews.

### Inclusion and exclusion criteria

The eligibility criteria were developed following discussions among the authorship team, which includes researchers with experience in the implementation of digital health in RRR settings and implementation science. These criteria were refined after being piloted on an initial set of reviews. The focus and boundaries of the search strategy are defined using the PICOS approach (population, intervention, comparator, outcome, study design) (Table 1).

**Table 1:** Inclusion and exclusion criteria.

| **Variable** | **Inclusion** | **Exclusion** |
| --- | --- | --- |
| Population | People who are the consumers for whom the DHI is designed to benefit, or people who deliver the innovation, or people who are the key-decision makers within the implementing setting. | No limitations. |
| Intervention | DHIs that focus on changing one or more health outcomes, quality of care, health, and care needs, or designed to support clinical decision making.<br>Technology as the main digital innovation or mode of delivery, may be supplemented with other forms of delivery. | Any use of a DHI solely for: (1) administrative purposes; (2) clinical education, training/coaching, or supervision; (3) app design or development, content analyses, adherence, or other technical details.<br>Studies predominantly about the general perceptions of digital health.<br>Studies in which the DHI is only a non-essential component of a more complex programme or intervention. |
| Comparison | No limitation. | No limitations. |
| Outcome | Primary outcome data on factors that influence the implementation of the DHI. | Studies focused on the measurement, development, testing or selection of digital health. |
| Study Setting | Studies focused in RRR Australia.<br>Settings may include, but are not | Studies not located in RRR areas and/or in Australia. |
|  | limited to acute care, outpatient care, long-term care, home-based care, and primary care settings.<br>Any clinical area, health, or quality of care outcome. |  |
| Study Type | Research on factors that influence implementation of DHI<br>Studies published in English<br>Studies published in a peer-reviewed journal | Studies where the influencing factors are not described in sufficient detail |

### Population

We will include reviews that comprise studies involving people (i.e., patients, healthcare providers, other end users) for whom the DHI is designed to benefit. This may include the people who are directly or indirectly involved with delivering the DHI, or key-decision makers who have the authority within the implementing setting.

### Intervention

We will include reviews that specifically examine DHIs that focus on transforming one or more health outcomes, quality of care, health service and healthcare needs, or clinical decision-making. The primary focus of these innovations is technology, serving as the main digital tool or mode of delivery, which may be complemented by other delivery methods. Studies will be excluded if they: (1) use technology solely for administrative purposes, clinical education, training/coaching, or supervision, (2) solely focus on design or development (i.e. DHI tools, apps etc.), content analyses, adherence, or technical aspects, or (3) are general perception, satisfaction or acceptance studies about digital health.

### Comparison

Reviews with or without comparative analyses of DHIs will be included. Where comparisons are made, these may include standard implementation processes, usual care, control groups, or alternative strategies.

### Outcome

Reviews that report on any objective or subjective influencing factor(s) to digital health implementation in RRR settings will be included. These may be detailed as barriers and/or enablers or reported as broader findings and reflections of the included literature (i.e., not categorised as barrier or enablers). Reviews specifically focused on the measurement, development, testing, or the selection of DHIs, will be excluded.

### Setting

We will include reviews that focus on studies in RRR Australia. This may include, but is not limited to healthcare environments, such as acute care settings (e.g., hospitals, emergency departments, oncology care centres, mental health facilities), outpatient care settings, long-term care settings (e.g., nursing homes, rehabilitation centres, palliative care settings, hospice care facilities), home-based care settings, and primary care settings (e.g., clinics, community health centres). Clinical area, or health / quality of care outcome is not restricted. To ensure we encompass all pertinent research, we will also include international reviews with at least one Australian-based study to be independently described at a country-level. Reviews will be excluded where a RRR setting are unable to be differentiated.

### Types of studies

We will include qualitative, quantitative, and mixed-methods systematic or scoping reviews with or without meta-analysis. Reviews must have identified literature by means of a structured search of bibliographic databases, have a transparent methodological criterion to exclude papers, and they must present critiquing conclusions related to the implementation of digital health. Primary research studies, rapid reviews, and efficacy/effectiveness reviews will be excluded. Reviews that are not available in English, where full text is unavailable, without a clear description of the review question, or eligibility criteria, or do not have evidence of critical appraisal, will be excluded. A PRISMA flow diagram will be used to summarise study selection.

### Screening and full-text review

First, all titles and abstracts will be independently examined by two independent reviewers (MK and SB), considering the pre-established inclusion and exclusion criteria. Inter-rater reliability will be established on 10% of the documents. Next, articles meeting the inclusion criteria will continue to the full-text screening phase, by the same two reviewers. Inter-rater reliability will be re-established to re-calibrate (if needed). Reasons for excluding articles from full-text review onwards will be recorded in the PRIOR flow diagram (28). Any disagreements will be resolved by consensus through discussion among the reviewers and a third reviewer (SL) will offer input if there are still unresolved conflicts. Reviewers will not take part in decisions about studies in which they were involved.

### Data extraction and management

Data will be extracted from all included reviews using a standardised form in an excel spreadsheet by one reviewer (MK) and cross-checked by another review member (LW) for accuracy. The data extraction template will be piloted using 10% of eligible full-text articles. The retrieval of data is in alignment with the research questions and is defined in Additional file 3. These data will be extracted only from included reviews.

### Managing overlapping studies

We acknowledge the risk of overlap in primary studies across multiple reviews, which can give undue weightage to studies included more than once in the synthesis of findings. We will assess the overlap in primary studies at the data extraction and synthesis stage. As recommended by Pieper et al. (29) we will create a citation matrix to visually demonstrate the amount of overlap and suitably address this in our analysis.

### Assessment of risk of bias in included studies

We will assess the methodological quality of included reviews using A MeaSurement Tool to Assess systematic Reviews (AMSTAR 2) rating tool (30). Studies with no or one non-critical weakness and no critical flaws will be rated as high quality; studies with more than one non-critical weakness and no critical weaknesses will be rated as moderate quality; studies with one critical flaw irrespective of non-critical weaknesses will be rated as low quality; and those with more than one critical flaw with or without a non-critical weakness will be rated as critically low quality. The domains considered critical are: (1) review methods established prior to conduct and any deviations reported; (2) comprehensive and reproducible literature search strategy; (3) providing justification for the exclusion of individual studies; (4) risk of bias assessment of the studies included in the review; (5) use of appropriate statistical methods in performing a meta-analysis; (6) accounting for risk of bias when interpreting the results; (7) evaluation of the presence and impact of publication bias. The AMSTAR 2 guidance states that reviews of critically low quality should not be relied on for comprehensive and accurate summaries of the literature. The risk of bias assessment will be conducted by one reviewer (MK) for all eligible reviews, and a 10% random selection by another reviewer (SB). Any discrepancies or disagreements will be resolved through discussion among these reviewers. The risk of bias assessments conducted for the primary studies in the included reviews will be narratively summarised.

### Data synthesis and evidence mapping

A narrative synthesis of the evidence will be conducted. The first step will include familiarisation with the extracted data through close reading, followed by coding. To ensure that coding remains grounded in the chosen theoretical framework, the publicly available CFIR codebook template will be used to develop an initial coding template specific to this study. The extracted data will be mapped to themes guided by the CFIR representing the determinants of implementation across five domains (innovation characteristics (i.e., strength of the evidence), outer setting (e.g., RRR context), inner setting (e.g., integrated delivery system), characteristics of individuals (e.g., healthcare professionals, service, participants) and implementation process (e.g., assessing context), and these will be categorised as barriers or enablers. Regular touchpoints will be held to discuss coding procedures among the team members involved.

We will construct an evidence and gap map defined as a visual depiction of the characteristics of evidence in a particular field (31). Our evidence map will highlight the characteristics of DHI research in RRR Australia, provide a broader contextual review of factors that influence implementation, and aid in emphasising the gaps in the research to inform the planning of future applications in RRR settings.

### Dissemination plans

The findings will be disseminated through a publication in a peer-reviewed journals in the field of implementation science, digital health, and rural or remote health. We will also disseminate results through relevant conferences, social media, and the Northern Australian Regional Digital Health Collaborative network (www.nardhc.org). Our evidence and gap map will be produced using an open-source platform, that will be interactive and publicly available. Furthermore, we will leverage existing connections through the national network of Advanced Health Research Translation Centres, leveraged by SL’s involvement to disseminate the findings of the review to a wider audience.

## Discussion

This overview of reviews will consolidate available evidence on the implementation of digital health in RRR Australia. Through our synthesis of quantitative, qualitative, and mixed-methods systematic and scoping reviews, we will answer two key questions: What factors inhibit or promote the implementation of digital health in RRR settings in Australia, and what implementation design constructs specific to RRR locations in Australia need to be considered? The attention to different settings and the inclusion of studies from RRR locations of Australia will provide a regionalised perspective, essential for understanding how context-specific factors may influence the generalisability of findings. This approach allows us to expose knowledge gaps, to orient further research that improves our understanding of the complex factors at play in the implementation of DHIs in RRR settings. For example, by applying mapping methods to existing taxonomies, implementation process strategies can be selected to match determinants across the CFIR levels of the individual, inner setting, and outer setting (32–35). This is a crucial step to designing tailored implementation strategies that are directly aligned with the specific barriers and enablers faced in RRR settings.

### Strengths and limitation

This review has several strengths. To enhance the review’s quality, we will adopt a predefined methodology based on the Cochrane Handbook for Systematic Reviews of Interventions.

Furthermore, we will assess the methodological quality of included reviews using the AMSTAR-2 checklist and apply a robust implementation framework (CFIR) to identify, categorise, and synthesise potential influences on implementation. Using CFIR will address a key limitation of current research in the field, since reviews and primary research are often focused on person-level barriers and enablers, omitting organisational- and system-level factors affecting digital health implementation.

The review protocol’s strength lies in its systematic and evidence-based approach. A limitation of this method is the restriction of findings to studies reported in systematic and scoping reviews, thereby excluding primary studies. We will minimise this risk by updating the search strategy at least once before the completion of the review. Another limitation is the potential overlap between primary studies. To address this, we will prepare a matrix of primary studies included in systematic reviews to avoid double counting of primary studies.

## Abbreviations

AMSTAR-2: A MeaSurement Tool to Assess systematic Reviews

CFIR: Consolidated Framework for Implementation Research

CINAHL: Cumulative Index to Nursing and Allied Health Literature

PICOS: Population, Intervention, Comparison, Outcomes and Study

PRIOR: Preferred Reporting Items for Overviews of Reviews

PROSPERO: International Prospective Register of Systematic Reviews

PRISMA-P: Preferred Reporting Items for Systematic review and Meta-Analysis Protocols

RRR: Rural, regional, and remote

## Declarations

### Ethics approval and consent to participate

Not applicable.

### Consent to publication

Not applicable.

### Availability of data and materials

All data generated or analysed during this study will be included in the published overview article.

### Author’s Contributions

MK and SL conceptualized the study. All authors were consulted throughout the protocol drafting process, provided a critical review of intellectual content and approved the final manuscript.

## Data Availability

All data produced from this study protocol will be available in a manuscript

## Acknowledgements

Not applicable.

## Additional Information

**Additional File 1.**
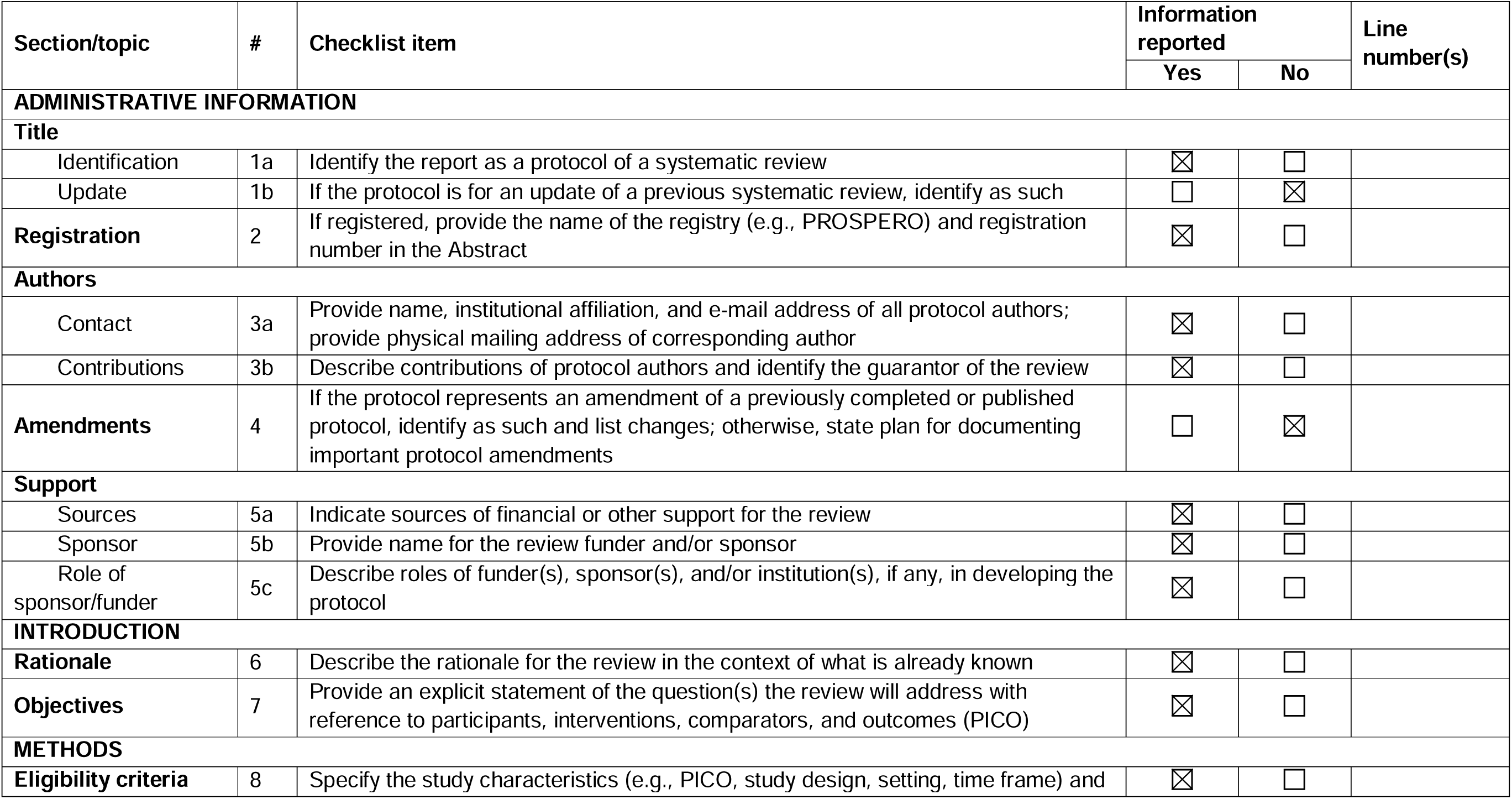
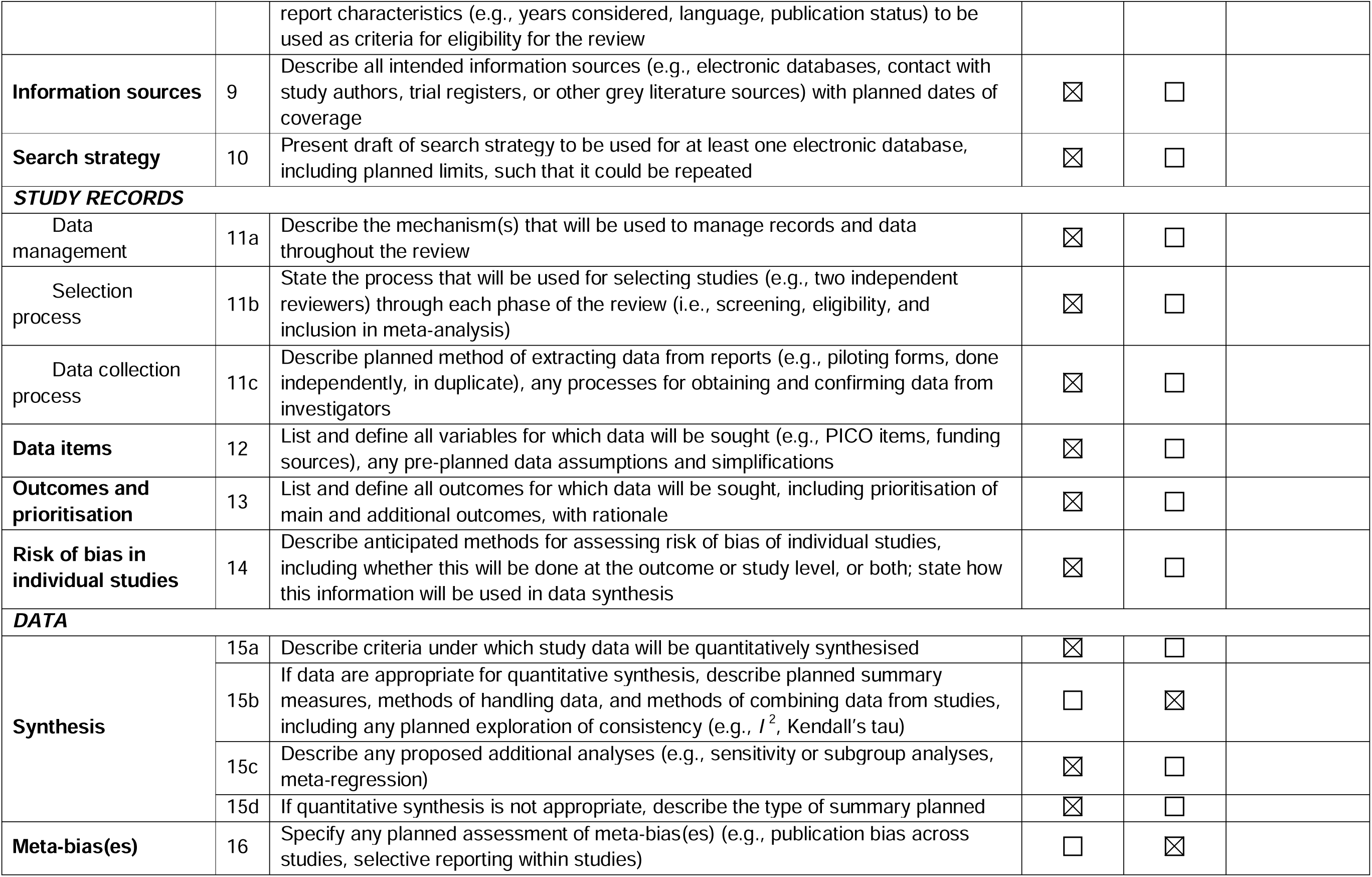
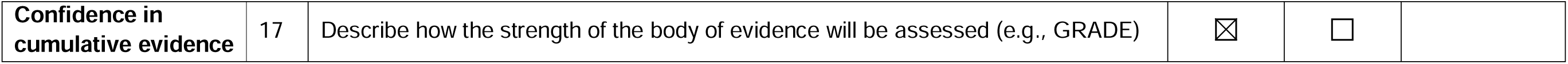
PRISMA-P 2015 statement.

| Section/topic | # | Checklist item | Information reported |  | Line number(s) |
| --- | --- | --- | --- | --- | --- |
|  |  |  | Yes | No |  |
| **ADMINISTRATIVE INFORMATION** |  |  |  |  |  |
| **Title** |  |  |  |  |  |
| Identification | 1a | Identify the report as a protocol of a systematic review | ☒ | ☐ |  |
| Update | 1b | If the protocol is for an update of a previous systematic review, identify as such | ☐ | ☒ |  |
| **Registration** | 2 | If registered, provide the name of the registry (e.g., PROSPERO) and registration number in the Abstract | ☒ | ☐ |  |
| **Authors** |  |  |  |  |  |
| Contact | 3a | Provide name, institutional affiliation, and e-mail address of all protocol authors; provide physical mailing address of corresponding author | ☒ | ☐ |  |
| Contributions | 3b | Describe contributions of protocol authors and identify the guarantor of the review | ☒ | ☐ |  |
| **Amendments** | 4 | If the protocol represents an amendment of a previously completed or published protocol, identify as such and list changes; otherwise, state plan for documenting important protocol amendments | ☐ | ☒ |  |
| **Support** |  |  |  |  |  |
| Sources | 5a | Indicate sources of financial or other support for the review | ☒ | ☐ |  |
| Sponsor | 5b | Provide name for the review funder and/or sponsor | ☒ | ☐ |  |
| Role of sponsor/funder | 5c | Describe roles of funder(s), sponsor(s), and/or institution(s), if any, in developing the protocol | ☒ | ☐ |  |
| **INTRODUCTION** |  |  |  |  |  |
| **Rationale** | 6 | Describe the rationale for the review in the context of what is already known | ☒ | ☐ |  |
| **Objectives** | 7 | Provide an explicit statement of the question(s) the review will address with reference to participants, interventions, comparators, and outcomes (PICO) | ☒ | ☐ |  |
| **METHODS** |  |  |  |  |  |
| **Eligibility criteria** | 8 | Specify the study characteristics (e.g., PICO, study design, setting, time frame) and report characteristics (e.g., years considered, language, publication status) to be used as criteria for eligibility for the review | ☒ | ☐ |  |
| **Information sources** | 9 | Describe all intended information sources (e.g., electronic databases, contact with study authors, trial registers, or other grey literature sources) with planned dates of coverage | ☒ | ☐ |  |
| **Search strategy** | 10 | Present draft of search strategy to be used for at least one electronic database, including planned limits, such that it could be repeated | ☒ | ☐ |  |
| **STUDY RECORDS** |  |  |  |  |  |
| Data management | 11a | Describe the mechanism(s) that will be used to manage records and data throughout the review | ☒ | ☐ |  |
| Selection process | 11b | State the process that will be used for selecting studies (e.g., two independent reviewers) through each phase of the review (i.e., screening, eligibility, and inclusion in meta-analysis) | ☒ | ☐ |  |
| Data collection process | 11c | Describe planned method of extracting data from reports (e.g., piloting forms, done independently, in duplicate), any processes for obtaining and confirming data from investigators | ☒ | ☐ |  |
| **Data items** | 12 | List and define all variables for which data will be sought (e.g., PICO items, funding sources), any pre-planned data assumptions and simplifications | ☒ | ☐ |  |
| **Outcomes and prioritisation** | 13 | List and define all outcomes for which data will be sought, including prioritisation of main and additional outcomes, with rationale | ☒ | ☐ |  |
| **Risk of bias in individual studies** | 14 | Describe anticipated methods for assessing risk of bias of individual studies, including whether this will be done at the outcome or study level, or both; state how this information will be used in data synthesis | ☒ | ☐ |  |
| **DATA** |  |  |  |  |  |
| **Synthesis** | 15a | Describe criteria under which study data will be quantitatively synthesised | ☒ | ☐ |  |
| | 15b | If data are appropriate for quantitative synthesis, describe planned summary measures, methods of handling data, and methods of combining data from studies, including any planned exploration of consistency (e.g., $I^2$, Kendall's tau) | ☐ | ☒ | |
|  | 15c | Describe any proposed additional analyses (e.g., sensitivity or subgroup analyses, meta-regression) | ☒ | ☐ |  |
|  | 15d | If quantitative synthesis is not appropriate, describe the type of summary planned | ☒ | ☐ |  |
| **Meta-bias(es)** | 16 | Specify any planned assessment of meta-bias(es) (e.g., publication bias across studies, selective reporting within studies) | ☐ | ☒ |  |

|  |  |  |  |  |
| --- | --- | --- | --- | --- |
| **Confidence in cumulative evidence** | 17 | Describe how the strength of the body of evidence will be assessed (e.g., GRADE) | ☒ | ☐ |

**Additional File 2.** Example of search strategy. Medline (PubMed) database

| Concept | Search string |
| --- | --- |
| #1 | (MeSH Terms ("Digital Health" OR "Precision Medicine" OR "Telemedicine" OR "Wearable Electronic Devices" OR "Medical Informatics") OR Title/Abstract ("Digital Health" OR "digital health care" OR "digital medicine*" OR "eHealth" OR "ehealth care" OR "e medicine*" OR "telehealth" OR "tele-health" OR "telehealthcare" OR "tele-healthcare" OR "Telemedicine" OR "tele-medicine" OR "mHealth" OR "m-health" OR "mHealthcare" OR "m-healthcare" OR "mobile health" OR "mobile healthcare" OR "mobile medicine" OR "online health" OR "online healthcare" OR "online medicine")) |
| #2 | MeSH Terms ("Rural Population" OR "Rural Nursing, Remote" OR "Rural Health Services" OR "Rural Health" OR "hospitals, rural") OR Title/Abstract ("rural" OR "remote communit*" OR "remote village*" OR "remote region*" OR "remote setting*" OR "regional area*" OR "regional communit*" OR "regional setting" OR "remote service*" OR "regional service*" OR "remote health" OR "regional health" OR "nonmetropolitan" OR "non metropolitan" OR "country area*" OR "country setting*" OR "remote area*" OR "rural and remote*") OR All Fields ("rural" AND "population") OR "rural population" OR ("rural" AND "nursing") OR "rural nursing" OR ("rural" AND "health" AND "services") OR "rural health services" OR ("rural" AND "health") OR "rural health") |
| #3 | Title/Abstract ("implement*" OR "innovat*" OR "integrat*" OR "uptake*" OR "up tak*" OR "accept*") OR All Fields ("scal*") AND Title/Abstract ("up" OR "adher*" OR "promot*" OR "adopt*" OR "engag*" OR "improvement*" OR "intervention*")) |
| #4 | Title/Abstract ("barrier*" OR "impede" OR "impediment*" OR "facilitat*" OR "challeng*" OR "solution*" OR "driver*" OR "optimi*" OR "diffus*" OR |
|  | "infus*" OR "success*" OR "limitation*" OR "strateg*" OR "approach*" OR<br>"hinder*" OR "obstacle*" OR "hurdle*" OR "opportunit*" OR "enabl*" OR<br>"sustain*" OR "operational*" OR "adapt*" OR "capacit*" OR "polic*") |
| #5 | #1 AND #2 AND #3 AND #4 |
| #6 | #5 AND (Australia*) |
| #7 | Limited to: (Review, Systematic Review [Publication Type] AND<br>English[Language]) |

**Additional File 3.** Characteristics for data extraction.

| Category | Information to be extracted |
| --- | --- |
| Article information | • Article title<br>• Authors(s)<br>• Year of publication<br>• DOI<br>• Journal |
| Objectives | • Primary objective/s<br>• Secondary objective/s |
| Methodological approach | • Databases searched<br>• Review period (searches / publications)<br>• Number of papers included (total / Australian)<br>• Number of primary studies and study designs<br>• Method of synthesis / analysis employed to synthesise the evidence |
| Population | • Participant characteristics |
| Innovation | • Technology type<br>• Mode of delivery<br>• Definition of digital health<br>• Intended purpose of the digital health |
| Setting or context | • Country<sup>a</sup><br>• Location in Australia (State/Territory)<br>• Rural, regional and/or remote<br>• Remoteness classification<sup>b</sup><br>• Setting details |
| Health focus | • Predominant health discipline/outcome<br>• Predominant health service or healthcare need<br>• Clinical decision-making<br>• Quality of care |
| Outcomes and review findings | • Outcomes reported by the author(s) relevant to the aims of this review:<br>○ Key themes and findings<br>○ Barriers and enablers<br>○ Lessons learned<br>○ Unintended consequences, risks, and implications |
| Evidence quality | • Instrument used to appraise quality/risk of bias of primary studies and the rating of their quality<sup>c</sup><br>• AMSTAR-2 rating |

<sup>a</sup> Refers to the location from all primary studies; <sup>b</sup>According to the Australian Standard Geographical Classification System - inner regional Australia, outer regional Australia, remote Australia, and very remote Australia; <sup>c</sup> It is possible that different tools, or distinct parts of tools to assess methodological quality/risk of bias have been used. In these situations, we will extract the disparate quality/risk of bias assessments to the best of our ability, despite the variability across reviews.

